# Red flags useful to screen for suspect cancer in patients with low back pain: a scoping review protocol

**DOI:** 10.1101/2024.09.01.24311072

**Authors:** Gianluca Notarangelo, Giuseppe Giovannico, Francesco Bruno, Claudia Milella, Firas Mourad, Filippo Maselli

## Abstract

Low back pain (LBP) is defined as pain and discomfort occurring under the costal arch and above the inferior gluteal folds, with or without leg pain. The most common type of low back pain is “non-specific low back pain”. It is defined as such because there is no specific pathology that can be the cause. Instead, “specific low back pain” is caused by diseases such as infections, tumors, osteoporosis, ankylosing spondylitis, fractures, inflammatory processes, radicular syndromes, cauda equina syndrome, etc. It is the most common musculoskeletal disorder in the world and the most cause of disability.

Most low back pain is benign in nature and specific diagnoses are uncommon; however in some specific cases this clinical presentation of signs and symptoms could hide much more serious conditions. The second most frequent serious pathology, after fracture, that may initially appears as low back pain is tumor pathology (0.2% and 7.0%), condition that turns out to be so dangerous as to be the second leading cause of death in the world according to the WHO.

Red flags are signs and symptoms that raise suspicion of the presence of serious diseases; these are clinical aspects of alert that can justify referral to the doctor/specialist and can contraindicate physiotherapy treatment.

Clinicians must therefore be able to recognize clinical presentations that are potentially dangerous to the patient that may require further evaluation or emergency referral. Failure to immediately recognize these conditions could lead to incorrect diagnoses resulting in worsening outcomes over time. The tools that will guide the clinician in recognizing the red flags in low back pain are a through medical history and a complete physical examination.

Currently we have very little clarity and agreement in the literature about the Red Flags that should be better investigated, a scoping review is strongly required and corresponded to the objectives of this project.

## BACKGROUND

Low back pain (LBP) is defined as pain and discomfort occurring under the costal arch and above the inferior gluteal folds, with or without leg pain. The most common type of low back pain is “non-specific low back pain”. It is defined as such because there is no specific pathology that can be the cause. Instead, “specific low back pain” is caused by diseases such as infections, tumors, osteoporosis, ankylosing spondylitis, fractures, inflammatory processes, radicular syndromes, cauda equina syndrome, etc. [1] It is the most common musculoskeletal disorder in the world and the most cause of disability [2-3].

After a correct and specific clinical evaluation, patients may suffer from the following conditions: specific spinal pathology (Vertebral fractures,Malignancy,Spinal infection, Axial spondyloartritis, Cauda equina syndrome;< 1% of cases in primary care), radicular syndrome (radicular pain, radiculopathy, spinal stenosis; 5-10% of cases in primary care) and non-specific LBP 90-95% [4]

Most low back pain is benign in nature and specific diagnoses are uncommon. [5]; however in some specific cases this clinical presentation of signs and symptoms could hide much more serious conditions. The second most frequent serious pathology, after fracture, that may initially appears as low back pain is tumor pathology (0.2% and 7.0%) [6], condition that turns out to be so dangerous as to be the second leading cause of death in the world according to the WHO.

Red flags are signs and symptoms that raise suspicion of the presence of serious diseases; these are clinical aspects of alert that can justify referral to the doctor/specialist and can contraindicate physiotherapy treatment. [7]

Clinicians must therefore be able to recognize clinical presentations that are potentially dangerous to the patient that may require further evaluation or emergency referral. Failure to immediately recognize these conditions could lead to incorrect diagnoses resulting in worsening outcomes over time. The tools that will guide the clinician in recognizing the red flags in low back pain are a through medical history and a complete physical examination. [8]

A recent study confirms that the presence of one or more Red Flags of low or intermediate diagnostic impact authorizes the clinician to “wait and see”, given that serious damage to the patient occurs after a delayed diagnosis of 4-6 weeks. Instead, the presence of Red Flag with a high diagnostic impact justifies a more thorough and urgent investigation and a probable referral to a spinal specialist. [9]

In addition to being a widely used correct model in the past, Red Flags screening has become a professional responsibility of the physiotherapist. It should be noted that the predictive power of this procedure still shows weaknesses and may not yet be sufficient in the field of Differential Diagnosis. [10]

IIn particular, this scoping review aims to:

1. Systematically map and summarize the current literature to identify any studies that reported RFs for cancer among patients who reported LBP;
2. Possibility of identifying a cluster of Red Flags that could help detect the presence of cancer.
3. Identify gaps in the evidence base and direct future research in this area.

## REVIEW QUESTION

The following research question was formulated: “What is known from the existing literature about RFs for cancer in patients presenting with LBP?

## METHODS

This scoping review will be conducted in accordance with the latest review process proposed by the Joanna Briggs Institute (JBI) in 2020 [11].

For the reporting, the Preferred Reporting Items for Systematic Reviews and Meta Analyses extension for Scoping Reviews (PRISMA ScR) [12] checklist will be used.

### Inclusion criteria

Full-text articles will be eligible for inclusion if they meet the following population, concept, and context (PCC) criteria:

- Population. This review will consider studies that included patients of any age and gender with any cancer diagnosis with reported LBP.
- Concept. This review will consider studies that explored and reported RFs for cancer among the population described above.
- Context. This review will consider studies conducted in any context.
- Sources. This scoping review will consider only studies studies published in English or Italian language. Publication date was restricted from 01 January 1999 to nowadays. No others geographical or setting restrictions will be applied.

### Exclusion criteria

Studies that do not meet the above-stated PCC criteria will be excluded.

### Search strategy

Literature research will be carried out on the following databases up to August, 15st 2024: MEDLINE, Scopus, Google Scholar Web of Science, Cochrane Library and SciELO.

The full search strategy for MEDLINE is available in the Appendix 1. We will check the reference lists of all identified studies.

### Study selection

Study selection Search results were collected and imported into EndNote V.X9 (Clarivate Analytics, PA, USA). Duplicates were automatically removed. The review process was conducted by two independent authors and consisted of two levels of screening using Rayyan QCRI online software [13]: (1) title and abstract review, and (2) full-text review. In case of disagreement, discrepancies were resolved by a third author. Reasons for excluding full-text sources were recorded and provided in the scoping review report. The results of the search will be reported in full in the final scoping review and presented in a Preferred Reporting Items for Systematic Reviews and Meta-analyses (PRISMA) flowchart.

### Data extraction

A standardized and planned Excel form will be used to sort the included studies and the extracted data. This extraction form will be created according to the PCC model and will be completed alternately by two authors, cross-checking each entry. A draft of the extraction tool is included in Appendix 2.

In each study examined, the extracted data will be: Author(s), Year of publication, Study design, Population, Characteristics, Sample size, Diagnosis of Low back Pain, Concept, RFs identified, Context, Country, Setting

The creation of charts of results is usually an iterative process in scoping reviews. Additional data may be added to this form depending on possible subgroups resulting from the analysis of the included studies. The changes are detailed in the full scoping review.

### Data synthesis

As a scoping review, the aim of this study is to summarize the results and provide an overview of the research rather than assessing the quality of individual studies [11]. The results are presented in two ways:

1. Numerical: We summarize and report the collected data as a descriptive analysis. We map the data and show the distribution of studies by publication period, study design and topic. Results are reported in table and summary format.
2. Thematic: A thematic summary on cancer RFs is performed. Additional descriptive subgroup analyses are reported (e.g., patient gender, lumbar spine disease).

## Data Availability

All data produced in the present work are contained in the manuscript.

## RELEVANCE AND DISSEMINATION

The results of this scoping review may increase clinicians’ knowledge regarding the identification of RF suspicious for cancer in patients with LBP and, consequently, suggest prompt referral. Overall, this review may provide relevant information that can help improve clinical care.

## AUTHORS’ CONTRIBUTIONS

All authors conceived, designed, drafted and approved the final protocol.

## Appendix 1: The full search strategy for MEDLINE Search conducted on 15^th^ of August 2024

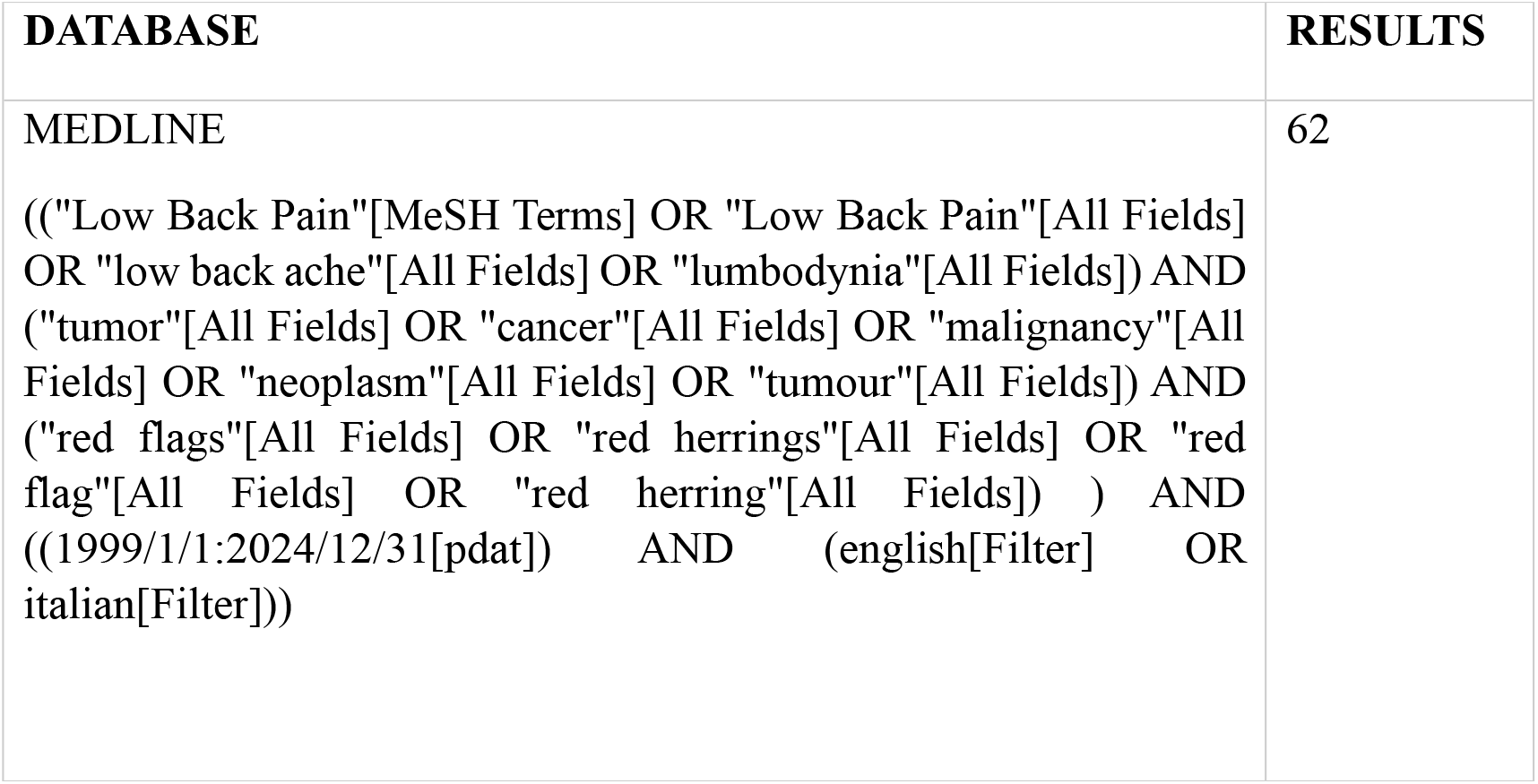

## Appendix 2: draft of the extraction tool

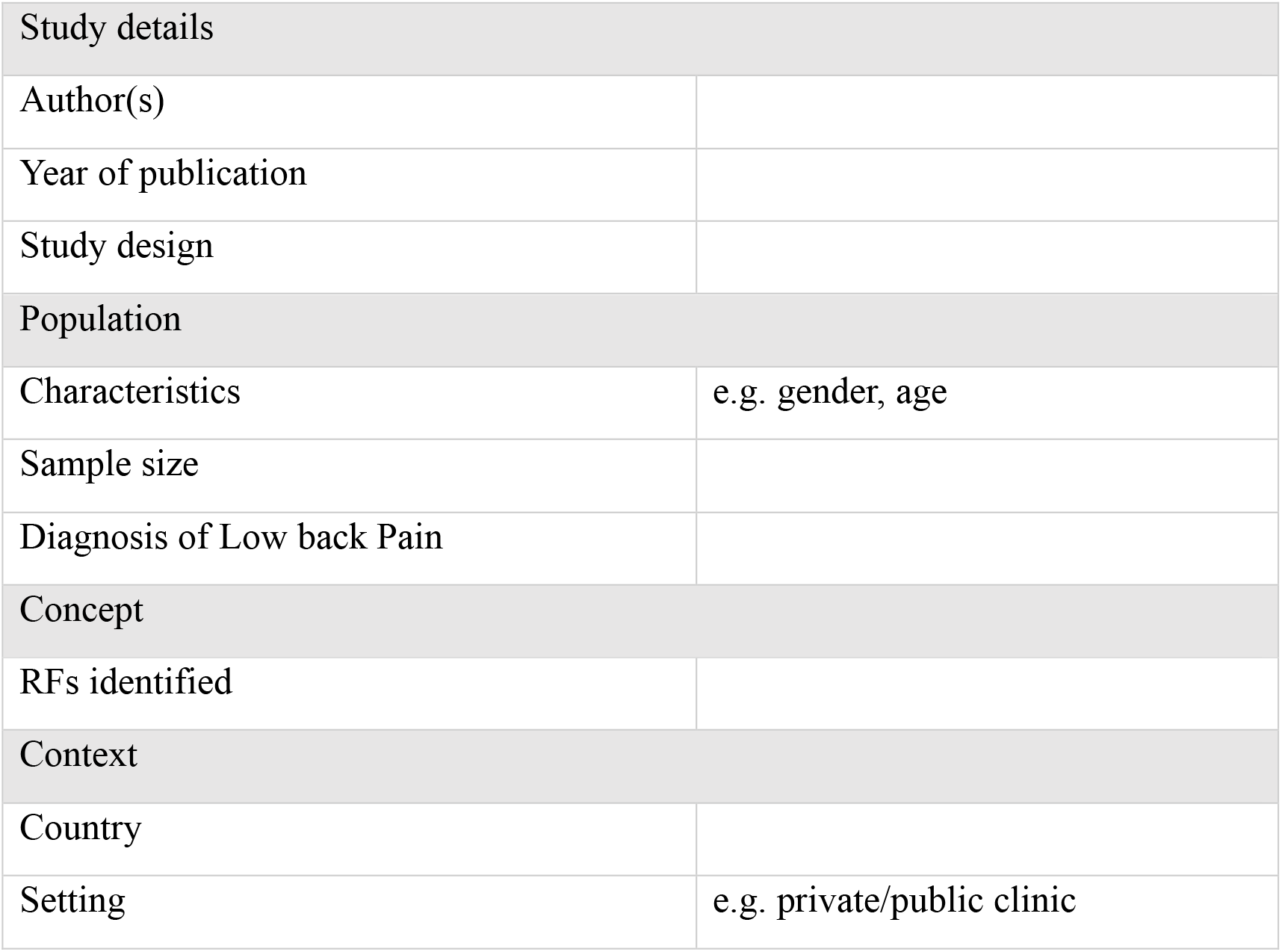

